# Perturbation of the Gut Microbiome and Association with Outcomes Following Autologous Stem Cell Transplantation in Patients with Multiple Myeloma

**DOI:** 10.1101/2022.05.19.22275261

**Authors:** Christopher D’Angelo, Sailendharan Sudakaran, Fotis Asimakopoulos, Peiman Hematti, Dalia El-Gamal, Nasia Safdar, Natalie Callander

## Abstract

The gut microbiome is an important feature of host immunity with associations to hematologic malignancies and cellular therapy. We evaluated the gut microbiome and dietary intake in patients with multiple myeloma undergoing autologous stem cell transplantation. Thirty patients were enrolled and samples were collected at 4 timepoints: pre-transplant, engraftment, day +100 (D+100), and 9-12 months post-transplant. Microbiome analysis demonstrated a loss of alpha diversity at the engraftment timepoint driven by decreases in *Blautia, Ruminococcus*, and *Faecalibacterium* genera and related to intravenous antibiotic exposure. Higher fiber intake was associated with increased relative abundance of *Blautia* at the pre-transplant timepoint. Lower alpha diversity at engraftment was associated with a partial response to therapy compared to complete response (CR) or very good partial response (VGPR) (CR/VGPR vs. PR, *p* <0.05). We conclude that loss of bacterial diversity at engraftment may be associated with impaired response to stem cell transplantation in multiple myeloma.

## INTRODUCTION

The prognosis for multiple myeloma patients has improved over the past two decades in part due to the use of autologous stem cell transplantation (ASCT) and lenalidomide maintenance therapy.[1] These treatments carry significant risks of toxicity including infection, diarrhea, and even death. Although responses can last for several years, a subset of patients have a more limited response, and relapse in all patients is an inevitable outcome in this incurable malignancy.[2]

The gut microbiome, a complex and abundant microbial community comprised of bacteria, archaea, viruses, protozoa, and fungi, is emerging as an important mediator of the host immune system.[3] Through incompletely understood mechanisms, the gut microbiome may influence tumor progression or response to therapies in many hematologic malignancies.[4,5] In allogeneic stem cell transplantation, higher gut microbiome diversity at time of cell engraftment has been linked to reductions in bacterial infections, graft-versus-host-disease, and even improvements in overall survival.[6-8] The presence of *Eubacterium* species has separately been associated with a higher degree of minimal residual disease (MRD) negativity in multiple myeloma and reduced risk of myeloma relapse after allogeneic stem cell transplantation.[7,9] In ASCT, a loss of bacterial diversity has been linked to reduced progression-free and overall survival.[10] Other studies have reported that the loss of bacterial diversity during ASCT may be driven by antibiotic exposure.[11,12]

Although the mechanism linking the microbiome to outcomes after ASCT remains unclear, intriguing work by Viaud et al demonstrated that cyclophosphamide, an alkylating agent with a similar mechanism of action to melphalan used in ASCT conditioning for myeloma, requires gut microbiota to obtain its optimal efficacy.[13] In a murine multiple myeloma model, the presence of *Prevotella heparinolytica* in the gut microbiome was linked to multiple myeloma progression through stimulation of IL-17 producing lymphocyte migration between the gut lumen and myeloma microenvironment.[14] A separate, potentially complementary mechanism implicating gut microbial species with nitrogen recycling pathways to enhanced glutamine synthesis may also lead to myeloma progression.[15]

As the associations between the gut microbiome and outcomes in multiple myeloma continue to emerge, it is increasingly important to validate and extend these findings given the multitude of potential confounding factors that may exist. For example, during ASCT, the use of different supportive care protocols, microbial sampling methods, antibiotic exposure, or conditioning regimens could inadvertently influence microbial populations. Additionally, the gut microbiome is highly influenced by diet but dietary analysis is typically not included in microbiome studies.[16] Furthermore, additional characterization of the factors driving the loss of microbial diversity will help understand how to transform the gut microbiome from a prognostic to a predictive biomarker and manipulate the microbiome-myeloma axis for therapeutic effect. To this end, we report on the results of a prospective study of the gut microbiome including dietary intake in a homogenous population of patients with multiple myeloma undergoing ASCT.

## METHODS

The study received Institutional Review Board approval. Informed consent was obtained from all subjects prior to any study conduct.

### Patient population and selection

Thirty subjects aged 18 years or older with a pathologically confirmed diagnosis of multiple myeloma undergoing ASCT with melphalan conditioning and planned lenalidomide/pomalidomide maintenance were consecutively recruited at the University of Wisconsin Carbone Cancer Center from 2019-2020. Subjects were excluded if they had a history of chronic diarrheal illness or inflammatory bowel disease, amyloidosis, non-measurable disease, or were anticipated to receive non-lenalidomide/pomalidomide maintenance at time of recruitment. Subjects who ultimately received a different maintenance therapy following enrollment could remain on study.

### Standard ASCT procedures

All subjects received melphalan 200mg/m^2^ conditioning 2 days prior to administration of a peripheral blood-collected autologous stem cell infusion featuring a minimum of 2.00 x10^6^ CD34+ cells/Kg ideal body weight without further graft manipulation. All subjects were hospitalized at time of stem cell infusion and discharged pending resolution of any acute transplant toxicities and evidence of stem cell engraftment, typically after 14 days. All subjects received acyclovir for varicella zoster prophylaxis on hospital admission. Fluconazole antimicrobial prophylaxis was added beginning at onset of neutropenia defined as an absolute neutrophil count (ANC)<1000 cells/μL and continued until the first day of neutrophil engraftment defined as an ANC >500 cells/μL. Levofloxacin antibiotic prophylaxis was started on the first day of neutropenia (ANC <1000 cells/μL) and continued until escalation of antibiotics for systemic infection or neutropenic fever or first day of neutrophil engraftment (ANC >500 cells/ μL), whichever occurred first. All subjects received G-CSF at 5 ug/Kg daily beginning day 5 till an absolute neutrophil count above 500/ μL. Neutropenic fever was treated empirically in all patients with vancomycin and cefepime and continued until ANC >500 cells/ μL or therapy could be specified based on culture data.

### Microbiome Sampling Procedures and Timeline

Subjects provided a first stool sample collected at home, ranging from 7 to 2 days prior to melphalan chemotherapy. Two subjects submitted a sample within 48 hours after stem cell infusion and were considered together in the first timepoint as these were collected prior to any changes in bowel habits or antibiotic exposure. A second sample was collected in the hospital post-transplant at the first day of neutrophil engraftment determined by an ANC >500 cells/μL. A third sample was collected approximately 100 days (D+100) post-transplant at the time of post-transplant response assessment and restaging. A fourth and final sample was collected 9-12 months post-transplant. All samples were immediately refrigerated at 4°C and returned to the lab for sample aliquoting into cryovials without additives or solutions and stored long-term at -80°C within 48 hours of collection.

### Diet History Survey Collection and Analysis

Subjects completed a dietary survey using the Diet History Questionnaire version 3.0 (DHQ-3) at the time of transplant and again at the D+100 restaging visit timepoint.[17] Briefly, this questionnaire was accessed and completed online using a computer, and subjects self-reported foods, beverages, and supplements taken in the last month, including how often these items were consumed. Self-reported dietary intake is then analyzed according to a database of 24-hour recall reporting from National Health and Nutrition Examination Surveys (NHANES) through DHQ-3 software to provide a mean daily intake of macro- and micronutrients. This data was compared at the two timepoints and analyzed using a Wilcoxon rank-sum test in R.

### Disease Assessment and Restaging

Disease assessment and response to therapy occurred pre-transplant and at the D+100 timepoint according to standard IWMG criteria for response assessment.[18] MRD testing was performed in a subset of patients using the 2-tube 8 color flow cytometry Euro-Flow methodology as described by Flores-Montera.[19]

### Microbiome DNA Processing and Sequencing

Approximately 80-100 mg of thawed stool was homogenized using glass beads and lysed using a cocktail of sodium dodecyl sulfate, lysozyme, mutanolysin, and lystostaphin.[20] Following enzymatic lysis, samples underwent a mechanical lysis step using bead-beating prior to phenol-chloroform DNA extraction.[20] Samples underwent DNA cleanup using a DNA Nucleospin cleanup kit per manufactuer’s directions (Macherey-Nagel, Germany) prior to library preparation and 16S rRNA gene sequencing using primers directed toward the V4 variable region and performed on the Illumina Miseq sequencer using established methods.[21]

### Microbiome Statistical Analysis

The resultant microbiome data were analysed using Quantitative Insights into Microbial Ecology (QIIME2) version 2.[22] Sequencing reads were denoised and quality filtered using the program DADA2.[23] The resultant sequence variants, equivalent to operational taxonomic units, were aligned and masked using MAFFT and the phylogenetic tree of the amplicon sequence variants was created using FastTree.[24,25] Taxonomy was assigned using a Bayesian classifier – Scikit-learn based on a pretrained Silva database (version 138) curated to the exact 16S amplicon region.[26] Alpha rarefaction curves using Shannon and Simpson indices were calculated for all samples. A rarefaction upper limit of median depth/sample count and the alpha diversity between different treatments was compared using Wilcoxon rank sum test. Samples were removed from further characterization if they did not contain sufficient reads. Beta-diversity was calculated and ordination plots generated using Bray–Curtis and Jaccard indices (non-phylogenetic) and weighted and unweighted UniFrac (phylogenetic) on amplicon sequence variant data leveled, according to the lowest sample depth. Permutation multivariate analysis of variance (PERMANOVA) testing was used for statistical analysis of beta-diversity measurements.

## RESULTS

### Baseline Demographics

Table 1 depicts the baseline disease and demographics for the enrolled subjects. Thirty subjects were recruited with a median age of 64 years (range 46-75) and an even distribution of males/females. The majority of subjects received standard triplet induction therapy typically comprised of bortezomib, lenalidomide, and dexamethasone. Nearly all subjects achieved a partial response prior to proceeding to transplantation.

**Table 1:** Patient Baseline Characteristics.

| Characteristic | Total n=30 (%) |
| --- | --- |
| Median Age (range) | 64 (46-75) |
| Sex |  |
| Male | 14 (47) |
| Female | 16 (53) |
| Disease Subtype |  |
| IgG | 17 (57) |
| IgA | 7 (23) |
| Light Chain | 6 (20) |
| Kappa | 23 (77) |
| Lambda | 7 (23) |
| R-ISS Stage* |  |
| I | 7 (23) |
| II | 18 (60) |
| III | 3 (10) |
| Pre-Transplant Disease Status |  |
| CR | 3 (10) |
| VGPR | 8 (27) |
| PR | 16 (53) |
| <PR | 3 (10) |
| Induction treatment |  |
| VRD | 23 (77) |
| VCD | 3 (10) |
| KRD | 3 (10) |
| RD | 1 (3) |

R-ISS: revised international staging system, CR: complete response, VGPR: very good partial response, PR: partial response, VRD: bortezomib, lenalidomide, dexamethasone, VCD: bortezomib, cyclophosphamide, dexamethasone, KRD: carfilzomib, lenalidomide, dexamethasone, RD: lenalidomide, dexamethasone. \* indicates 2 subjects had insufficient information for R-ISS staging.

### Nutrition Evaluation

Prior to transplant, patients reported an average calorie (1830 kcal/day), protein (77 g/day), and fruit intake (1.9 cups/day) that approximates recommendations from the United States Department of Agriculture (USDA) guidelines.[27] Alternatively, patients were notably deficient in fiber (17g/day), vegetable (1.7 cups/day), and whole grain (0.78 cups/day) prior to transplant. Following transplant, when re-assessed at the D+100 timepoint, patients lost an average of 5% of pre-transplant body weight. Additional modest changes between pre- and post-transplant dietary evaluation include a modest decline in mean calorie intake (1830 cal/day vs 1510, *p* = 0.04) and dietary fiber (17g vs 14g, *p* =0.03). No differences were observed in vegetable or fruit intake. Subjects overall diet was graded using the Healthy Eating Index (HEI), which is determined by how each subjects food intake conforms to dietary recommendations provided by the USDA and HHS.[28] This score is measured from 0-100, where 100 indicates perfect adherence to guideline recommendations and 0 indicates no adherence. For comparison, the average HEI for Americans is 59/100. Overall, the average HEI for subjects did not differ significantly between the two timepoints (median average and interquartile range 61 (54-69) vs 63 (58-70) *p* = 0.5, pre- and post-transplant, respectively) and approximates the score for average Americans.

### Gut Microbiome Analysis

Overall, 113 of 120 planned samples (4/subject, 30 subjects total) were collected through the study course. On average, we observed relative high microbial diversity prior to transplant (Figure 1), which dropped considerably immediately after transplant at time of neutrophil engraftment before recovering to pre-transplant levels by D+100 and onward. Beta diversity analysis, which compares differences between two communities, revealed a degree of overlap but overall distinctive microbial populations between samples collected pre transplant and post-transplant at time of engraftment, which were statistically significant by PERMANOVA testing (*p* = 0.01), suggesting that different microbial communities were formed after transplant.

**Figure 1:**
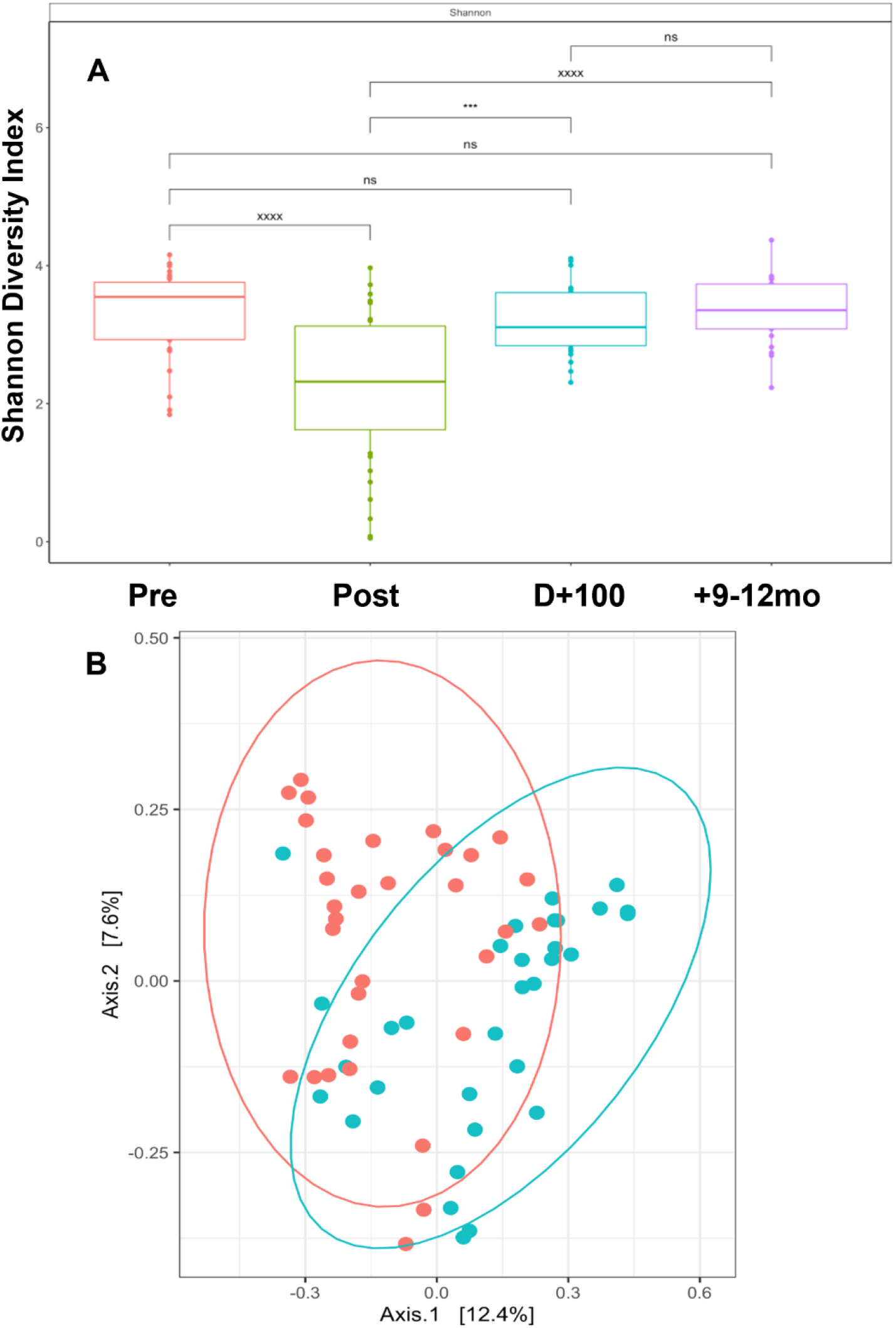
Gut Microbiome Diversity Changes Over Time. **A:** Shannon alpha diversity index according to 4 timepoints: Pre (pre-transplant), Post (first day of neutrophil engraftment), D+100 (100 day post-transplant), +9-12mo (9-12 months post transplant. *** refers to p-value <0.005, xxxx refers to p-value <0.0005. NS: non-significant. **B:** Beta-diversity assessment using Bray-Curtis metric. Red dots are pre-transplant samples, Teal dots are post-transplant samples at neutrophil engraftment.

To determine which taxa were differentially present between samples, we performed Lefse analysis to identify the taxa associated with the largest changes between the pre-transplant period and post-transplant engraftment timepoint.[29] Figure 2A depicts the cladogram describing this data. The loss of diversity was largely driven by loss of *Clostridial* species, particularly members of the lachnospiraceae family. The top genera lost post-transplant included *Blautia* species, *Ruminococcus* species, and *Faecalibacterium* species (Figure 2B). Although there were overall few gains in taxa post-transplant, members of the *Lactobacillus* genus, *Veillonella* genus, and *Escherichia_Shigella* genus demonstrated the largest increases in abundance post-transplant (Supplemental Figure 1).

**Figure 2:**
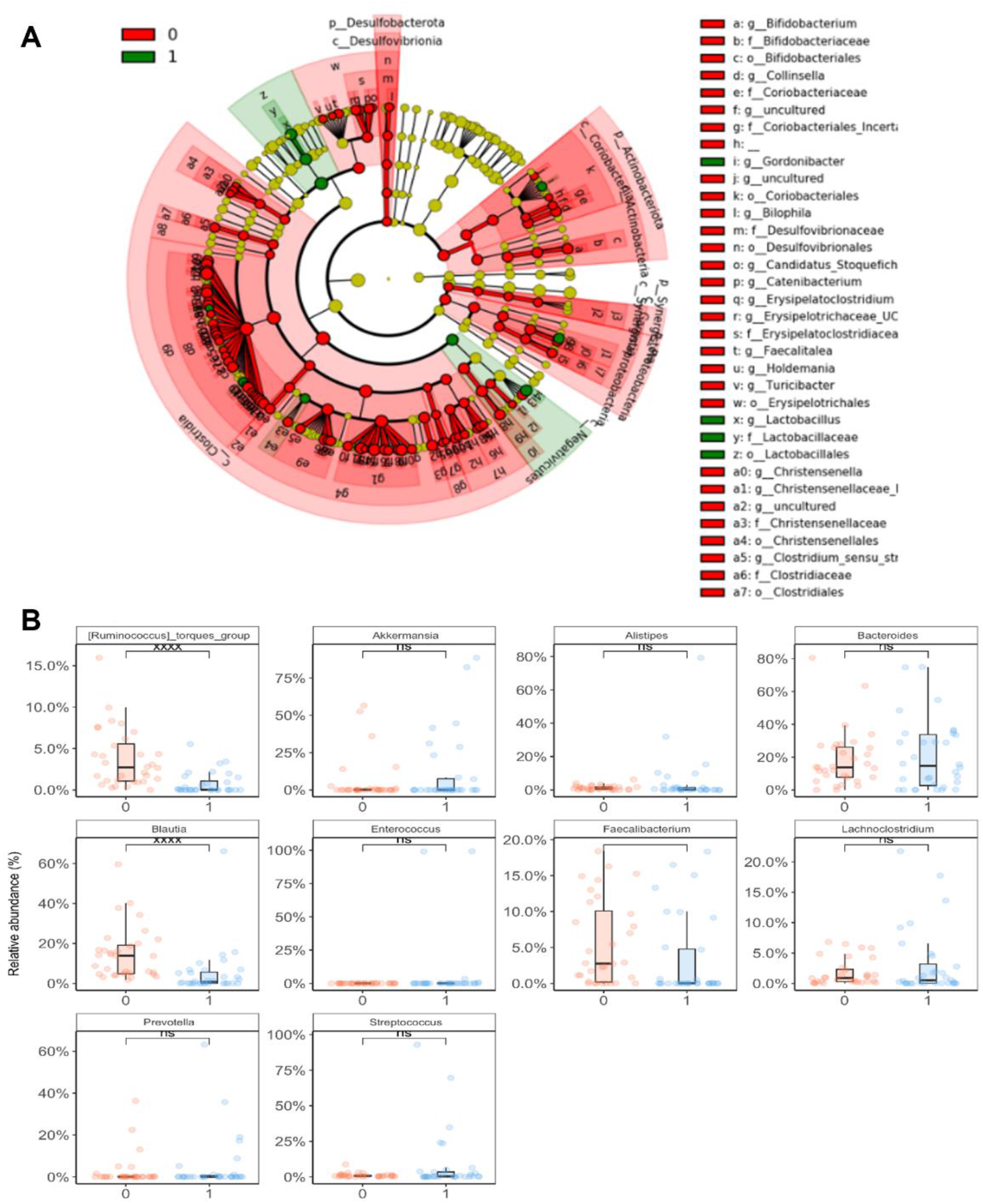
Relative Taxa Changes Pre/Post Transplant. **A:** Cladogram depicting taxa changes between pre transplant (red, timepoint 0) and immediate post-transplant at time of neutrophil engraftment (green, timepoint 1). Taxa depicted in red are relatively increased in abundance pre-transplant. Taxa in green are increased in post-transplant samples. **B:** Relative abundance changes in top 10 abundant genera between pre-transplant (red, timepoint 0) and post-transplant engraftment (blue, timepoint 1) samples. NS: non-significant, xxxx: p-value <0.0005. Ruminococcus, Blautia, and Faecalibacterium demonstrated a statistically significant difference in abundance between timepoints.

We hypothesized that pre-transplant fiber intake and overall healthy eating as measured by the HEI may influence gut microbiota community structure. To test this, we separated fiber intake and the HEI into quartiles and examined differences in pre and post-transplant alpha and beta-diversity metrics. Overall, there were no clear differences in fiber quartile and alpha diversity (Figure 3A). Although we observed the greatest pre-transplant alpha diversity in the highest fiber quartile, this relationship was not statistically significant when compared to the lowest fiber quartile. Similarly, no relationship between the HEI and alpha diversity was observed (data not shown). When evaluating taxa specific differences according to fiber intake (Figure 3B), we observed a dose-dependent increase in the *Blautia* genus according to higher fiber intake at the pre-transplant timepoint. A step-wise decrease in the *Bacteroides* genera was observed by increasing fiber quartile as well. No other clear trends in other top taxa and fiber intake in the pre-transplant timepoint were observed.

**Figure 3:**
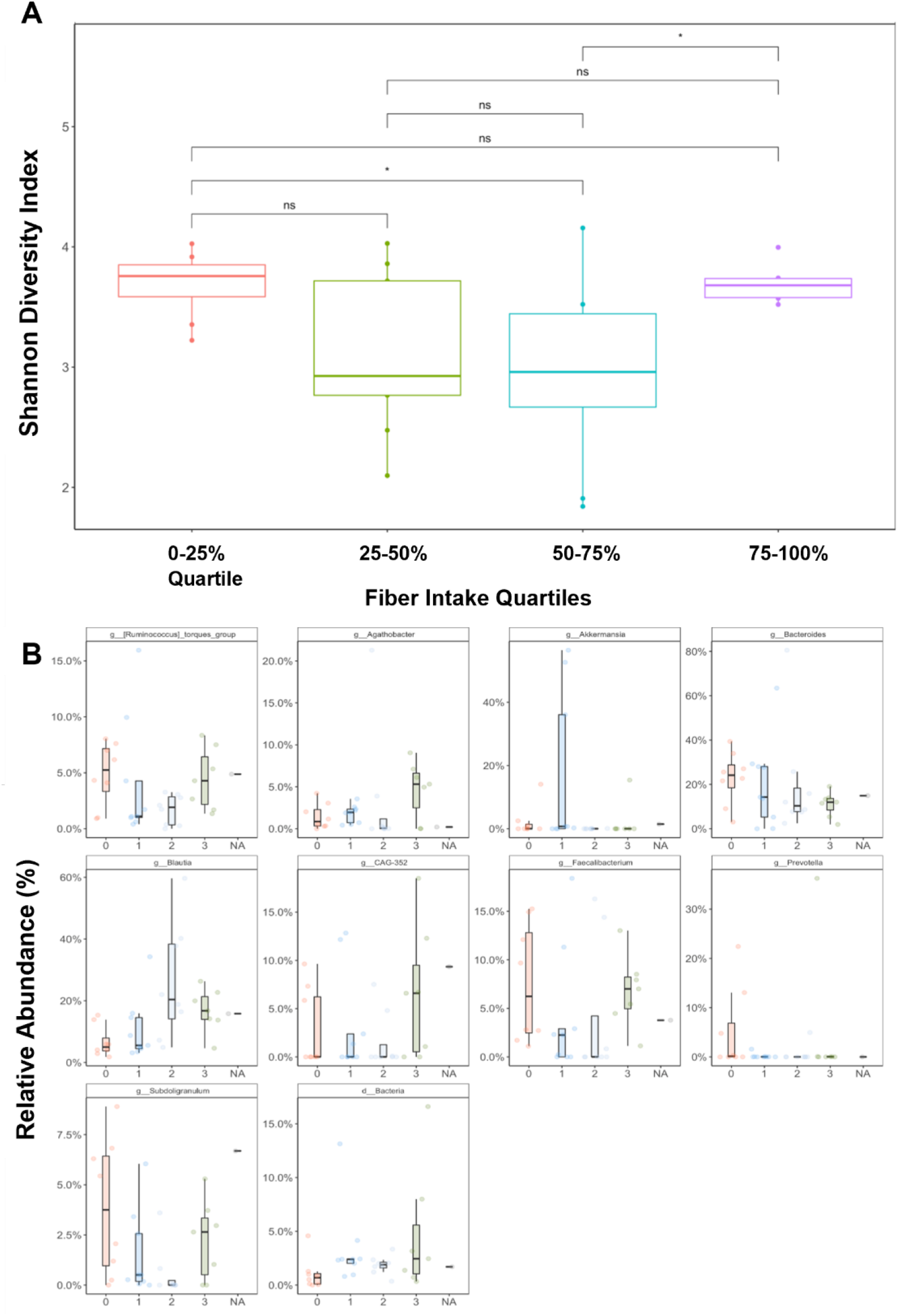
Effect of Fiber Intake on Gut Microbiome Community Structure. **A:** Boxplot depiction of Shannon alpha diversity index at pre-transplant timepoint according to fiber intake quartile. NS: not statistically significant, * *p* <0.05 **B:** Relative abundance changes in top 10 abundant genera between fiber quartiles (0: 0-25%, 1: 25-50%, 2: 50-75%, 3: 75-100%) at pre-transplant timepoint.

### Impact of Antibiotic Exposure on Post-transplant Diversity

In exploring the main drivers of loss of microbial diversity post-transplant, we observed that several subjects had particularly little change in diversity between the pre- and post-transplant timepoints (data not shown). Review of clinical data identified a key difference in IV antibiotic exposure, which was confirmed when examining post-transplant diversity changes by IV antibiotic exposure (Figure 4A). Twenty-four subjects developed a neutropenic fever (24/30, 80%) and received IV antibiotics while 6 subjects (6/30, 20%) did not, receiving only levofloxacin as antimicrobial prophylaxis during transplant. Using Lefse analysis, we identified the top taxa changes and note that the largest loss in taxa relative abundance following exposure to IV antibiotics were in the *Blautia and Faecalibacterium* genera (Figure 4B).

**Figure 4:**
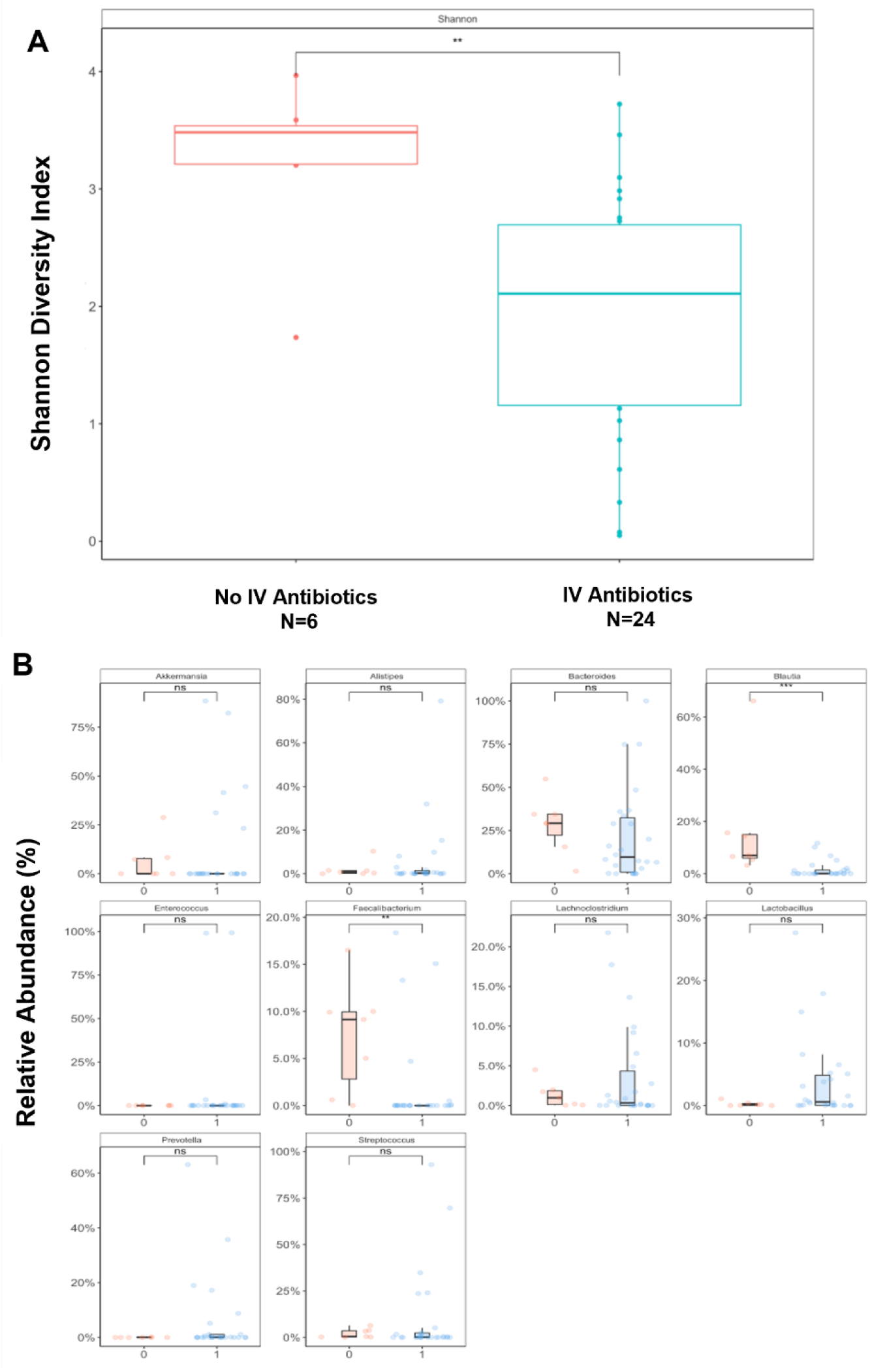
Effect of Antibiotic Exposure on Post-Transplant Diversity. **A:** Boxplot depiction of Shannon alpha diversity index at post-transplant engraftment timepoint according to IV antibiotic exposure. ** *p* value <0.01 **B:** Relative abundance changes in top 10 abundant genera between antibiotic exposure at post-transplant timepoint. 0: no IV antibiotics, 1: IV antibiotic exposure. NS: not significant. ** *p* value <0.01, *** *p* value <0.005

### Relationship of Gut Microbiota and Post-Transplant Response

We examined the relationship between alpha diversity at engraftment and response to transplant at D+100. Out of 29 evaluable subjects (1 subject withdrew from the study), there were 11 complete responses (CR, 38%), 10 very good partial responses (VGPR, 35%), 6 partial responses (PR, 21), and 2 with progressive disease (PD, 7%). We observed a statistically significant association between higher alpha diversity and CR or VGPR versus PR, which was associated with a lower degree of alpha diversity (*p* < 0.05 CR/VGPR vs PR, Figure 5). Although changes in the relative abundance of the *Blautia* genus were noted, these were not statistically significant and no predominant taxa change was identified (Supplemental Figure 2). Similarly, we evaluated the relationship between MRD positivity and the gut microbiome at engraftment. There were no differences in alpha diversity, but an increase in *Bacteroides* species was noted in the MRD positive versus MRD negative subjects (*p* < 0.05, Supplementary Figure 3).

**Figure 5.**
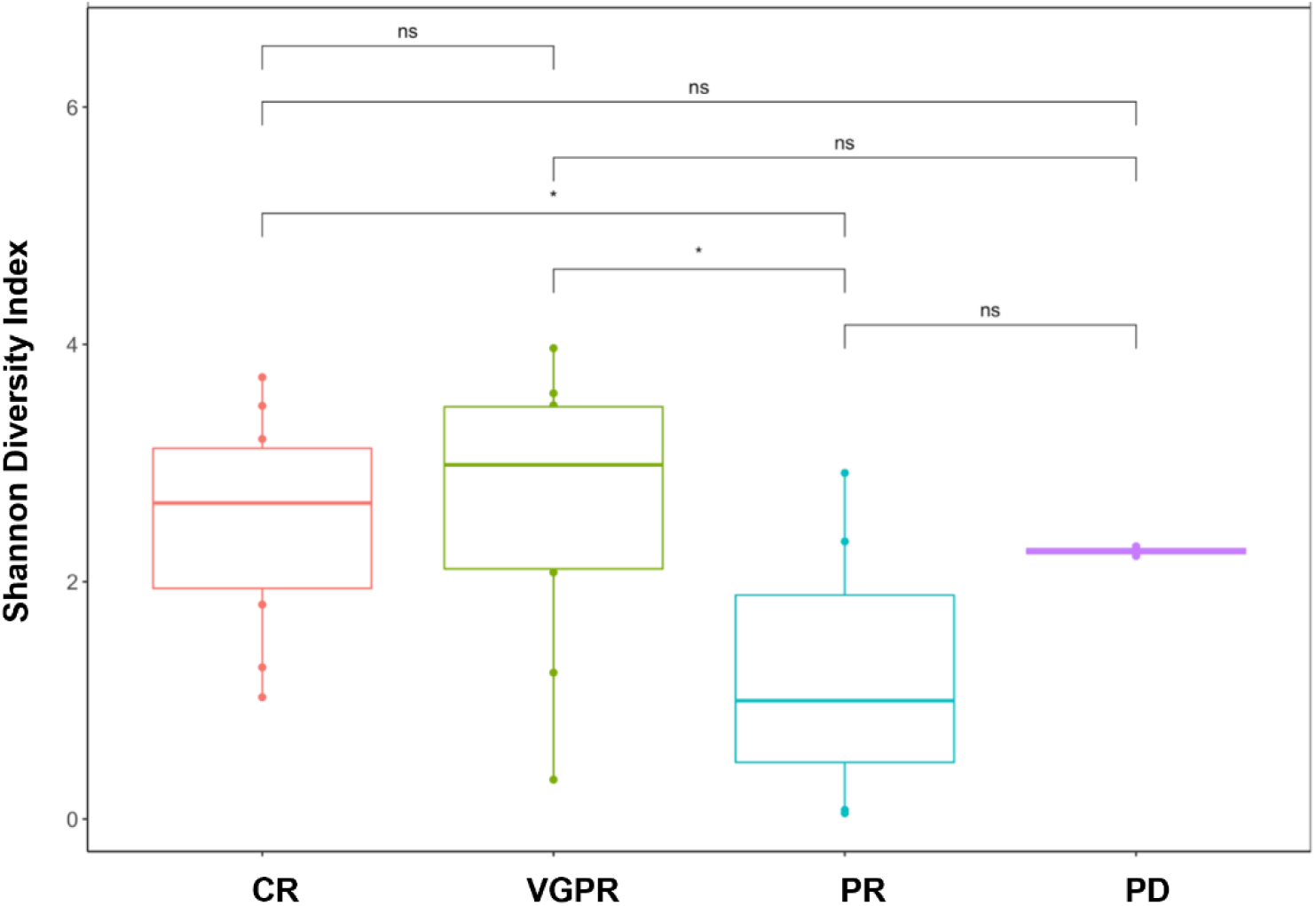
Association of Gut Microbial Diversity at Engraftment and Response. **A:** Boxplot depiction of Shannon alpha diversity index according to clinical response to transplant at D+100 assessment. CR: complete response, VGPR: very good partial response, PR: partial response, PD: progressive disease. NS: not significant, * *p* <0.05

## DISCUSSION

We performed a prospective evaluation of dietary intake and microbiome changes in the first year following ASCT in patients with multiple myeloma. Our analysis reveals several key findings. First, we confirm the results of prior published work suggesting a major loss of bacterial diversity in the immediate-post transplantation period.[10,11] We extend that work by identifying that the diversity of the gut microbiome is relatively re-established by D+100 post-transplant and maintained at the 9-12 month post-transplant period after the induction of maintenance therapy. We confirm that microbiota changes in the immediate post-transplant period are largely driven by IV antibiotic exposure, irrespective of the use of antibiotics with or without anaerobic activity. Antibiotic exposure is increasingly recognized to cause gut microbiome disturbances, with evidence suggesting an association to impaired survival outcomes in both cellular therapy and immunotherapies including immune checkpoint inhibitors, most recently demonstrated in Hodgkin lymphoma.[11,30,31] We identified that changes observed in alpha diversity between the pre-transplant and engraftment timepoints are largely driven by the loss of species in the *Clostridia* class, specifically members of the *Lachnospiraceae* family and *Blautia, Ruminococcus*, and *Faecalibacterium* genera. These species have independently been associated with improved outcomes either after allogeneic stem cell transplantation, or in the context of immunotherapies including chimeric antigen receptor therapy or programmed death-1 inhibition.[31-34] Given their potential importance in both cellular and immunotherapy in oncologic disease, these findings merit further investigation into both their association with clinical outcomes after cellular therapy and mechanism by which they may influence these outcomes.

The finding that alpha diversity is associated with response to ASCT provides a key link to prior published work suggesting that higher alpha diversity is associated with improved progression-free and overall survival in multiple myeloma.[10] These results strengthen the overall association between alpha diversity and clinical outcomes following ASCT, suggesting that alpha diversity of the gut microbiome may serve as a prognostic biomarker. Although the biologic mechanism governing this relationship requires further elucidation, a relationship between peri-transplant antibiotic exposure, loss of microbial diversity, and deleterious effects on transplant outcomes is emerging. Importantly, melphalan is an alkylator therapy whose mechanism of action is similar to cyclophosphamide. Work by Viaud et al and Iida et al have demonstrated that antibiotic ablation of the gut microbiome impairs the efficacy of cyclophosphamide in tumor-bearing mouse models.[13,35] These observations may indicate a similar mechanism is occurring for melphalan chemotherapy, where deleterious antibiotic effects on the gut microbiome in the immediate post-transplant period may impact the efficacy of melphalan therapy. Melphalan exposure frequently leads to gastrointestinal mucositis, where changes to the gut barrier may allow for microbial translocation events that could lead to activation of the immune system. This hypothesis is supported by studies conducted in mice depicting the influence of gut microbial intestinal translocation events on improving responses in an adoptive T-cell therapy via activation of the innate immune system.[36] Separately, evidence from Sivan et. al. supports that gut microbiome species influence dendritic cell activation leading to greater activation of anti-tumor T-cell responses in a mouse melanoma model.[37]

A high alpha diversity within the gut microbiome is generally associated with a healthy microbial community.[3] Loss of such diversity suggests a substantial impact on the community, yet it is unclear if high alpha diversity, and the community network it is representative of, is itself associated with optimal transplant outcomes. Alternatively, a high alpha diversity may simply be a marker for important bacterial taxa, where loss of diversity parallels the loss of key taxa. Future work to determine whether it is the bacteria taxa, the available genetic repertoire, or a metabolic niche responsible for these observations will be necessary to better understand this relationship.

To our knowledge, this is the first study to examine dietary changes and gut microbiota findings in patients undergoing ASCT. Patients with plasma cell disorders are increasingly interested in dietary counseling, but often do not receive counseling from their oncologists and rely on non-medical sources.[38]. We demonstrate that patients with myeloma undergoing transplant report dietary fiber intake pre-transplant (17g/day) that falls well below the recommended daily intake (28g/day) by the USDA.[27] Fiber intake provide key nutrients to beneficial gut microbial species and has been associated with reductions in certain solid cancers, and low fruit intake has been linked with myeloma progression.[39-42] Our findings may help counsel patients by providing a baseline on average pre-transplant dietary intake and identification of potential nutritional deficiencies. We note that baseline differences in fiber intake were associated with changes to microbial community structure, where a step-wise increase in the presence of the *Blautia* genus and decrease in *Bacteroides* genus was observed with increasing fiber intake. Given the beneficial role ascribed to *Blautia* in allogeneic stem cell transplantation, our findings provide support for further clinical trial investigations of dietary measures to modulate the microbiome as a means of influencing responses to ASCT in multiple myeloma.[32]

Our study is limited by its small sample size, reducing our ability to draw definitive conclusions in dietary associations to gut microbiota and ability to identify specific taxa relationships to clinical outcomes. In conclusion, we demonstrate that changes to the gut microbiome occur post-transplantation, are predominantly driven by IV antibiotic exposure, and that reduced alpha diversity at time of cell engraftment is associated with impaired responses to ASCT. These findings confirm and extend prior work in this field and provide further justification to both explore the gut microbiome as a biomarker and potential target of intervention in cellular therapy for multiple myeloma.

## Data Availability

All data produced in the present study are available upon reasonable request to the authors

## Acknowledgements

The authors thank the patients and their families for participating in this research project. The authors also thank the members of the Safdar lab including Ashley Kates, Lauren Watson, Nathan Putnam Buehler, Jared Godfrey, and Courtney Deblois for all their efforts in helping with sample collection and microbiome sequencing. Dr. D’Angelo was supported by a T32 training grant through the National Heart, Lung, and Blood Institute (5T32HL007899-20) and by a NIH loan repayment award through the National Cancer Institute (NCI). This work was additionally supported through a NIH/NCI R01CA252937 award to Dr. Asimakopoulos.

## Data Availability Statement

Data are available on request to the corresponding author.

## Figures and Tables

**Supplementary Figure 1:**
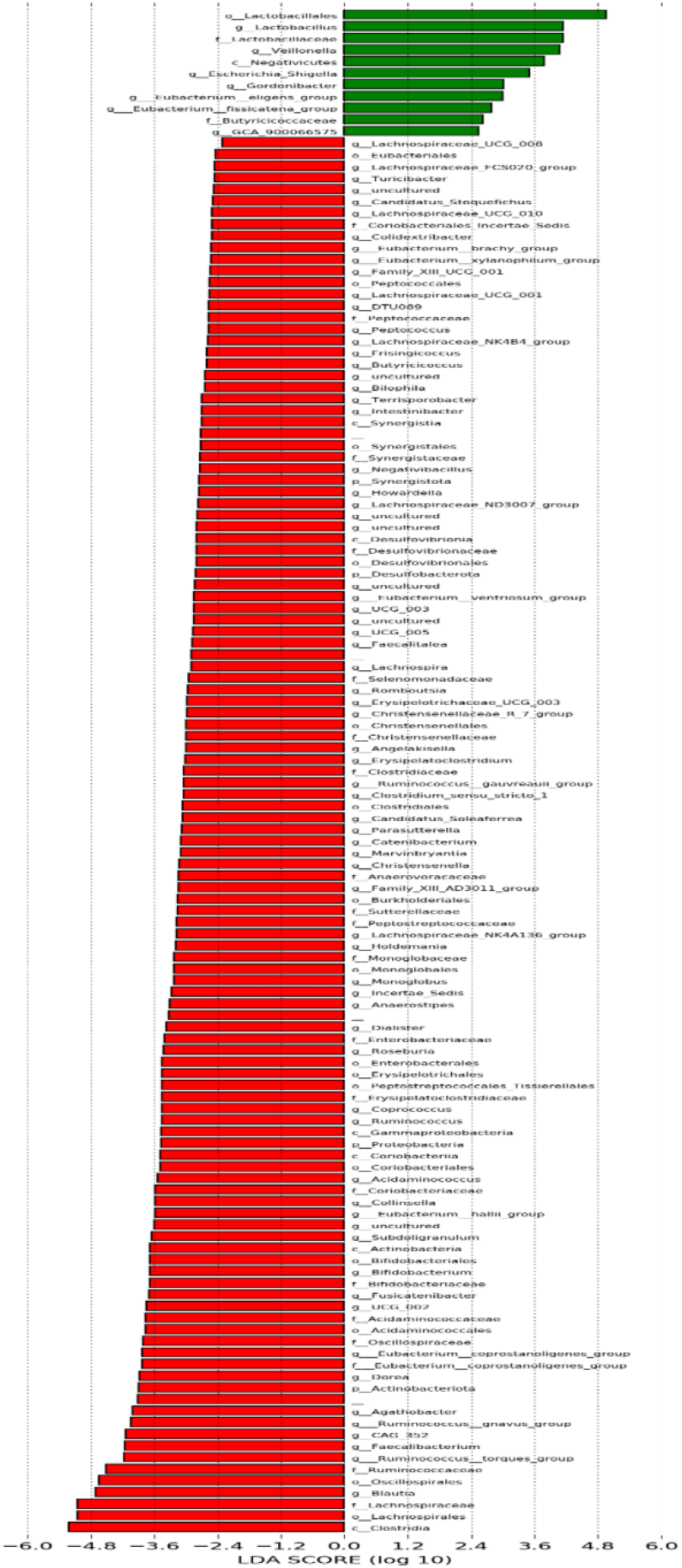
LEfSe Analysis of Top Taxa Changes Post Transplant. Red: Pre-transplant species with increased relative abundance. Green: taxa with relative increase in abundance at the post-transplant engraftment timepoint compared to pre transplant levels

**Supplementary Figure 2:**
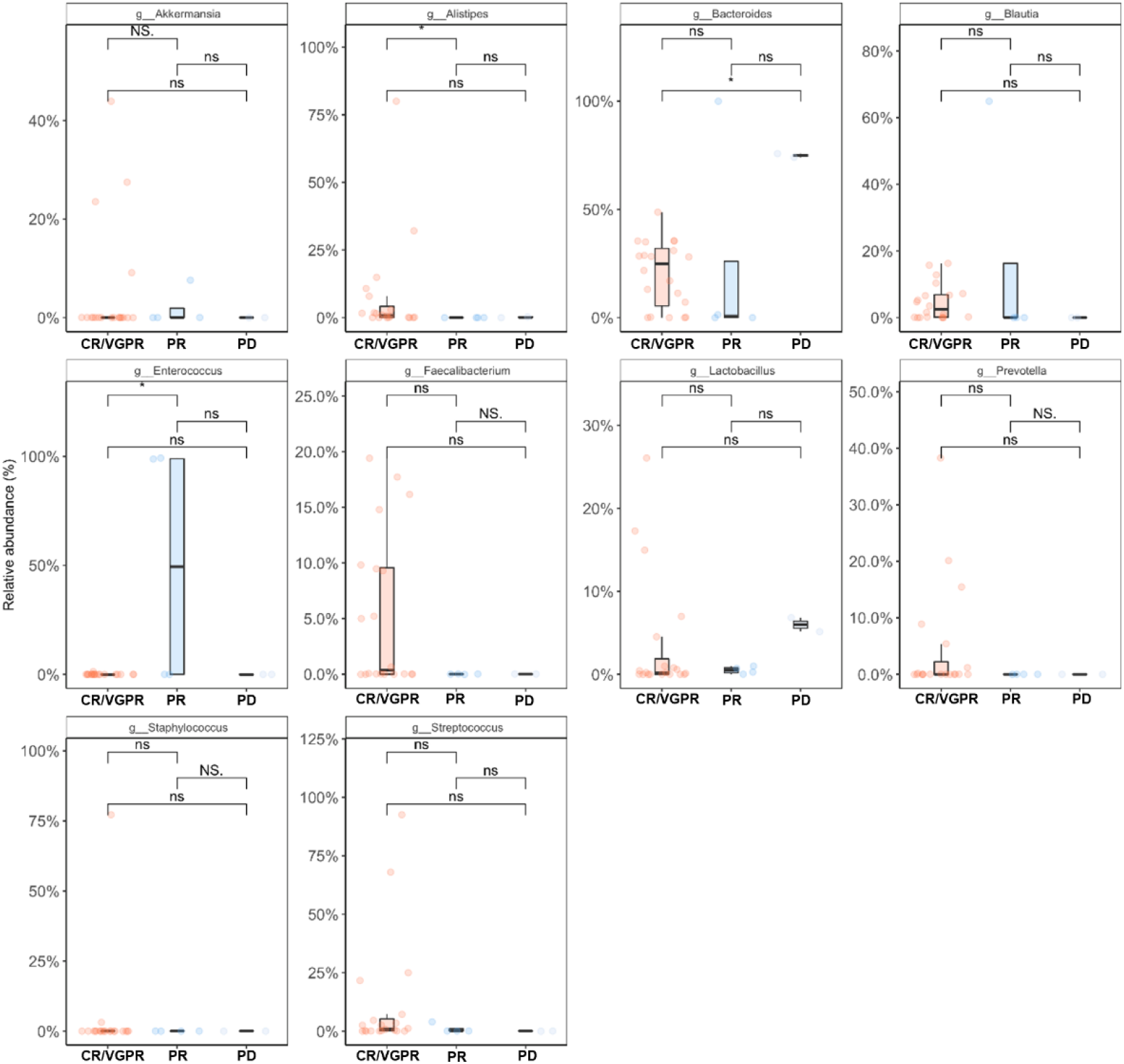
Relative Abundance Differences by Post-Transplant Response. Relative abundance changes in top 10 abundant genera in the post-transplant engraftment timepoint according to post-transplant response. CR: complete response, VGPR: very good partial response, PR: partial response, PD: progressive disease

**Supplementary Figure 3:**
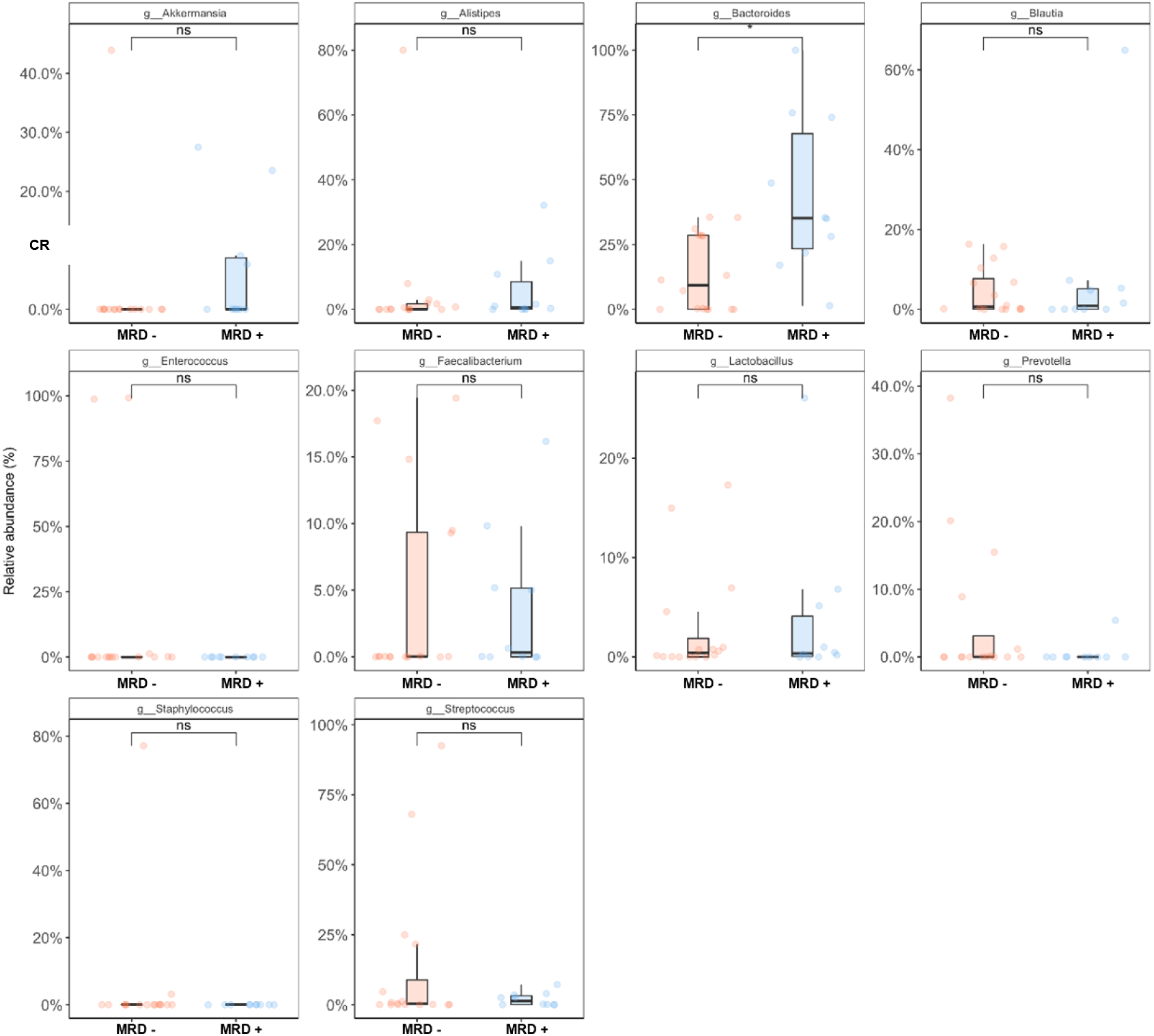
Relative Abundance Differences by MRD Status. Relative abundance changes in top 10 abundant genera in the post-transplant engraftment timepoint according to post-transplant MRD status. MRD-: minimal residual disease negative, MRD+: minimal residual disease positive

